# AI-driven Integration of Multimodal Imaging Pixel Data and Genome-wide Genotype Data Enhances Precision Health for Type 2 Diabetes: Insights from a Large-scale Biobank Study

**DOI:** 10.1101/2024.07.25.24310650

**Authors:** Yi-Jia Huang, Chun-houh Chen, Hsin-Chou Yang

## Abstract

The rising prevalence of Type 2 Diabetes (T2D) presents a critical global health challenge. Effective risk assessment and prevention strategies not only improve patient quality of life but also alleviate national healthcare expenditures. The integration of medical imaging and genetic data from extensive biobanks, driven by artificial intelligence (AI), is revolutionizing precision and smart health initiatives.

In this study, we applied these principles to T2D by analyzing medical images (abdominal ultrasonography and bone density scans) alongside whole-genome single nucleotide variations in 17,785 Han Chinese participants from the Taiwan Biobank. Rigorous data cleaning and preprocessing procedures were applied. Imaging analysis utilized densely connected convolutional neural networks, augmented by graph neural networks to account for intra-individual image dependencies, while genetic analysis employed Bayesian statistical learning to derive polygenic risk scores (PRS). These modalities were integrated through eXtreme Gradient Boosting (XGBoost), yielding several key findings.

First, pixel-based image analysis outperformed feature-centric image analysis in accuracy, automation, and cost efficiency. Second, multi-modality analysis significantly enhanced predictive accuracy compared to single-modality approaches. Third, this comprehensive approach, combining medical imaging, genetic, and demographic data, represents a promising frontier for fusion modeling, integrating AI and statistical learning techniques in disease risk assessment. Our model achieved an Area under the Receiver Operating Characteristic Curve (AUC) of 0.944, with an accuracy of 0.875, sensitivity of 0.882, specificity of 0.875, and a Youden index of 0.754. Additionally, the analysis revealed significant positive correlations between the multi-image risk score (MRS) and T2D, as well as between the PRS and T2D, identifying high-risk subgroups within the cohort.

This study pioneers the integration of multimodal imaging pixels and genome-wide genetic variation data for precise T2D risk assessment, advancing the understanding of precision and smart health.

## Introduction

Medical imaging has emerged as an indispensable auxiliary instrument for facilitating clinical diagnostics, playing a crucial role in various applications, such as breast cancer diagnosis using mammography (MMG) ^1^, fatty liver diagnosis through abdominal (ABD) ultrasonography ^2^, stroke diagnosis utilizing magnetic resonance imaging (MRI) and computed tomography (CT) ^3^, and carotid artery stenosis screening using carotid artery ultrasonography (CAU) ^4^.

Recently, artificial intelligence (AI) has revolutionized imaging analysis and its applications ^5, 6^. AI diagnostic models trained by deep learning neural networks, such as convolutional neural network (CNN) ^7^ and generative adversarial network (GAN) ^8^, based on different imaging-modality data, provide an automated and cost-benefit way to assist medical doctors in disease diagnosis and lesion detection. AI deep learning models have been developed for automated medical diagnostics for smart health, including fatty liver diagnosis based on ABD images ^9, 10, 11^, thyroid nodule detection based on thyroid ultrasound (TU) images ^12^, breast cancer diagnosis using MMG images^13^, diabetic retinopathy diagnosis using color fundus (CF) images ^14, 15^, atrial fibrillation and normal sinus rhythm detection using electrocardiogram (ECG) images ^16^, and osteoporosis diagnosis using bone mineral density radiography (BMD) images ^17, 18^.

In clinical practice, Imaging-Derived Features (IDFs) are obtained either through imaging technology utilizing automated measurement algorithms or through the expertise of medical technologists who strategically enhance medical imaging with manual or semi-manual annotations. These IDFs are crucial in helping medical doctors optimize disease diagnosis and select the Region of Interest (ROI). Influential IDFs can link medical imaging and diseases, constituting the primary imaging biomarkers for disease diagnosis and classification.

“**Fe**ature-**C**entric **A**nalysis (FECA)” and “**Pix**el-Based **A**nalysis (PIXA)” present two major imaging data analytical approaches. They employ distinct data and methodologies for disease diagnosis, presenting two contrasting approaches, each with its advantages and limitations. FECA leverages the IDFs derived from medical doctors’ annotations and observations, incorporating expert insights from medical technologists to enhance diagnostic accuracy. However, this approach significantly escalates the workload and cost of extracting annotations from medical images. Additionally, incorporating numerous IDFs may complicate data and variable collection, diminishing clinical applicability.

In contrast, PIXA eliminates the need for manual imaging labeling, offering an automated, low-labor, and cost-effective analysis. This approach is particularly beneficial for large-scale data processing. However, due to certain technical constraints, PIXA may overlook clinical expertise and contextual understanding^19^. This raises a practical question regarding the information richness and clinical plausibility of the two strategies in precision medicine: Can medical imaging inherently provide all requisite information, eliminating the need for human annotation by domain experts? In other words, is a hybrid approach, where data-driven methods are used initially and expert consultation is sought for final decision-making, recommended? Or does the knowledge of medical experts significantly contribute insights beyond medical imaging for disease diagnostics and classification, necessitating their inclusion at an early stage?

In addition to comparing PIXA vs. FECA, this study also compared multi-modality analysis (MUMA) vs. single-modality analysis (SIMA) in disease risk evaluation. MUMA allows for a more comprehensive assessment of the studied subject by integrating structural, functional, anatomical, and molecular information from multiple imaging modalities with increased sensitivity and specificity in disease diagnosis, classification, and subtyping, such as MRI and PET imaging, were combined to the classification of Alzheimer’s disease ^20, 21^. Integration of medical imaging and clinical features for breast cancer classification and subtyping ^22, 23^ and lung cancer classification and subtyping ^24^. Integrating these modalities provides a more holistic understanding of the etiology of the studied diseases under investigation. However, this approach may increase technical complexity, computational demand, and data acquisition cost compared to SIMA.

The convergence of medical imaging and genetic data within large-scale biobanks, driven by artificial intelligence and data sciences, marks a transformative paradigm shift in precision health for T2D. Our previous research aimed to consolidate IDFs from four distinct medical imaging modalities—abdominal ultrasonography (ABD), carotid artery ultrasonography (CAU), bone density scan (BMD), and electrocardiography (ECG)—alongside genome-wide single-nucleotide-polymorphism (SNP) data to assess T2D risk ^25^. This innovative analysis resulted in a high-accuracy risk evaluation model, polygenic risk score (PRS), and multi-imaging risk score (MRS), facilitating the identification of high-risk subgroups. Moreover, the model recommended eight crucial risk factors, including family history, age, fatty liver, spine thickness, PRS, end-diastolic velocity in the right common carotid artery, RR interval, and end-diastolic velocity in the left common carotid artery. The result highlights the importance of genetics and medical imaging in a precision medicine revolution.

Building on the previous work ^25^ based on a FECA, the current study concentrates explicitly on PIXA of ABD and BMD images alongside whole-genome SNPs for 17,785 Han Chinese participants from the Taiwan Biobank for T2D risk evaluation. The Taiwan Biobank, a national data repository from a Han Chinese population in Taiwan, aims to recruit 200 thousand participants with comprehensive data, including medical imaging, whole-genome genotyping, questionnaires, and lab tests ^26^. Given the predominant focus of many biobanks on European populations, the Taiwan Biobank stands out as a valuable resource for exploring medical imaging, genetic data, and precision medicine in East Asian populations ^27^.

This study uses CNN-based deep-learning models to analyze raw pixel data to generate a convolutional activation representation. Concurrently, using Bayesian statistical learning models, we analyze genome-wide SNP data to derive a PRS. Subsequently, the eXtreme Gradient Boosting (XGBoost) machine learning approach ^28^ integrates the imaging activation vector, genetic PRS, and demographic variables as classification and disease risk evaluation predictors. In imaging analysis, we also employ a graph neural network (GNN) to account for correlations among images within an individual and to integrate these multiple images into a unified representation. These advancements in PIXA and MUMA, coupled with genetic and demographic variable integration, present a promising avenue for developing fusion models encompassing deep learning, machine learning, and statistical learning in artificial intelligence and data sciences dedicated to disease risk evaluation. This contributes significantly to enhancing our understanding of precision health for T2D.

## Study participants and materials

### Participants

The study included 17,785 Han Chinese participants from the Taiwan Biobank, each of whom possessed both genetic and medical imaging data; the medical imaging included abdominal (ABD) imaging data (multiple-organ images) and bone mineral density (BMD) imaging data (spine, left hip, and right hip images). A participant was classified as a Type 2 Diabetes (T2D) case if they self-reported T2D, and had hemoglobin A1C (HbA1c) levels ≥6.5% or fasting glucose (GLU-AC) levels ≥126. A control was a participant who self-reported non-T2D, with HbA1C levels ≤5.6% and GLU-AC levels <100. These criteria included 7,786 participants consisting of 1,118 T2D cases and 6,668 non-T2D controls (**Fig. 1A**).

**Figure 1.**
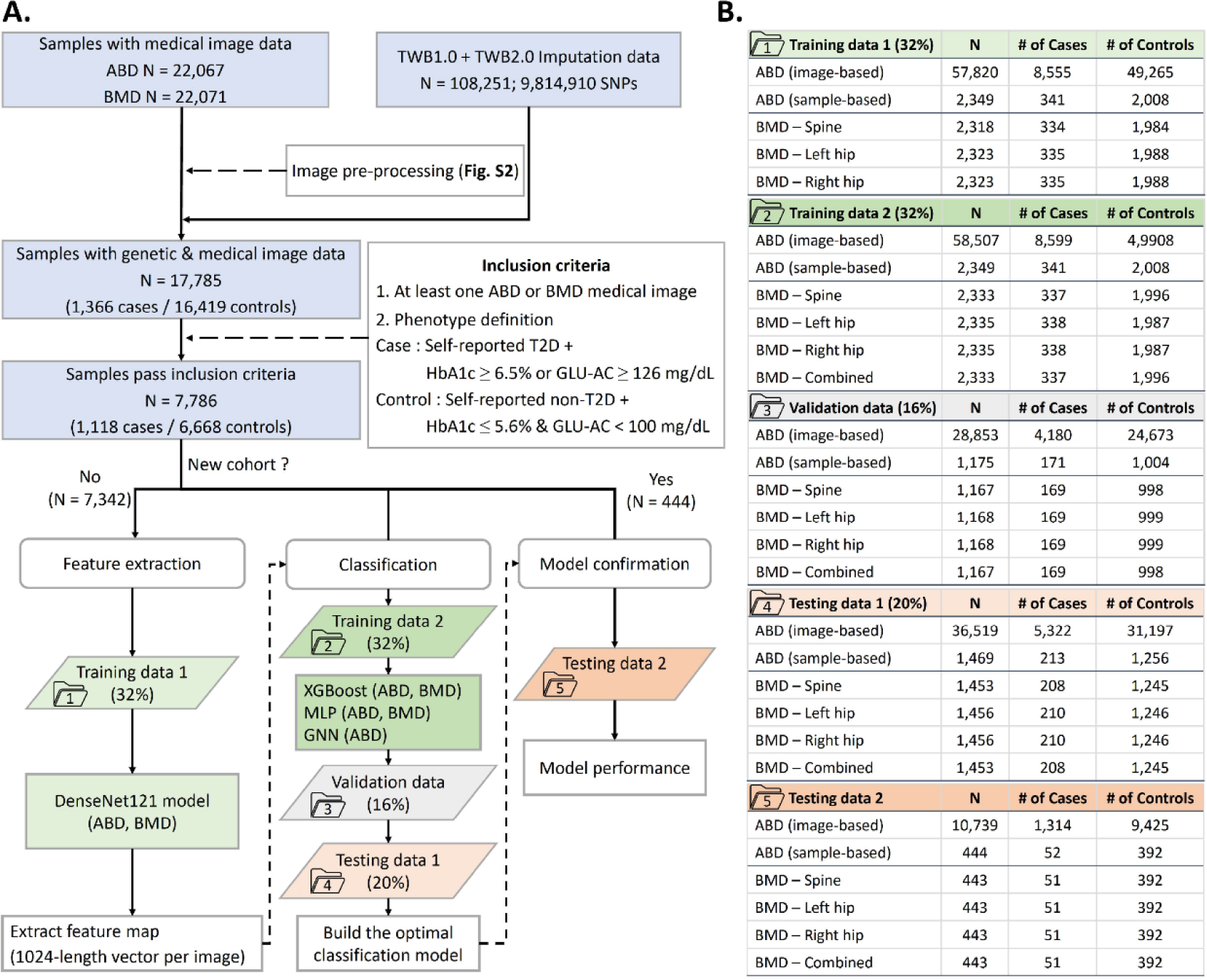
Flowchart of the study. **(A) Data extraction and classification model building.** In this dataset, 21,927 individuals possess ABD and BMD medical imaging data, while 108,251 individuals have whole-genome genotyping and imputation data. Moreover, 17,785 individuals possess both medical imaging and genetic profile data. Finally, the final dataset comprises 7,786 individuals who meet the inclusion criteria based on self-reported T2D status, HbA1C, and fasting glucose, consisting of 1,118 T2D cases and 6,668 normal controls. The complete dataset was initially divided into training + validation and testing sets at 8:2. Subsequently, the training + validation set was further separated into training and validation datasets with an 8:2 ratio. The training set was divided into two independent subsets at a 5:5 ratio to mitigate the winner’s curse problem; a two-stage procedure was employed for feature extraction and classification. In the first stage, which was dedicated to feature extraction, the first training data was used to establish a DenseNet121 model based on the initial training dataset (Training Dataset 1). Subsequently, a feature map vector was obtained. The second stage was focused on sample classification. Utilizing the data from the second training (Training Dataset 2) and validation datasets, a deep learning model for T2D classification was developed, and the results were further confirmed using the Testing Dataset 1. Ultimately, the best model’s validity was further confirmed using the second independent testing dataset (Testing Dataset 2) (n = 444). Regarding the deep learning classification model, three methods— multilayer perceptron (MLP), graph neural network (GNN), and eXtreme Gradient Boosting (XGBoost)—were implemented. **(B) Sample Size.** Information on the total sample size, number of cases, and number of controls is provided.

Among 7,786 participants, the dataset of 7,342 participants was collected earlier and initially divided into a training + validation set and testing set, named “Testing Dataset 1,” at an 8:2 ratio. Subsequently, the training + validation set was further divided into a training dataset and a validation dataset, named “Validation Dataset,” using an 8:2 ratio. Furthermore, the training set was randomly partitioned into two distinct subsets at a 5:5 ratio – the first subset, named “Training Dataset 1,” was utilized for training the feature extraction model, while the second subset, named “Training Dataset 2,” was employed to establish the classification model independently. Finally, an additional 444 participants were recruited later, thus regarded as a new cohort for the independent “Testing Dataset 2” (**Fig. 1B**).

### Demographic variables

Demographic variables were collected through questionnaires. The study incorporated age, sex, and family history of T2D, where the family history was quantified by the count of T2D cases among the father, mother, brother, and sister (ranging from 0 to 4). The epidemiological characteristics of these variables within the study population were detailed (**Table S1**).

### Image pixel files and image-derived features

For ABD medical imaging, each sample comprised multiple images depicting various organs. The imaging data encompassed the raw image file (DICOM format) (**Fig. S1**) and 28 image-derived features (IDFs) (**Table S1**). All the 28 IDFs were obtained through medical experts’ assessment. For BMD medical imaging, each sample included a single image for each type of BMD medical image, explicitly focusing on the spine, left hip, and right hip (**Fig. S1**). The imaging data for BMD included the raw image file (DICOM format) (**Fig. S1**) and 79 IDFs (**Table S1**). BMD machines automatically generated all 79 IDFs. Further details about the medical imaging protocol can be found on the Taiwan Biobank website (https://www.biobank.org.tw/english.php).

### Single nucleotide polymorphisms and imputation

All participants underwent genotyping at the National Center for Genomic Medicine at Academia Sinica using the Axiom TWB1.0 and TWB2.0 SNP arrays, comprising 653 thousand and 750 thousand SNPs, respectively. Additional information on the SNP annotation is available on the Taiwan Biobank website (https://www.biobank.org.tw/about_value.php).

Sample and marker quality controls followed the procedures in our previous study^25, 29^. Pre-phasing imputation was performed for TWB1.0 and TWB2.0 individually using SHAPEIT2 and IMPUTE2 (v2.3.1). The imputation process yields a probability distribution for each locus and each genotype of an individual. The PLINK command “--hard-call-threshold 0.1” was used to convert probabilities into actual genotypes, with interpretations made only when probabilities were greater than or equal to 0.9. If all three genotype probabilities fell below 0.9, the locus for that individual was considered missing. TWB1.0 and TWB2.0 imputation data were merged, and loci with missing rates exceeding 5% and minor allele frequency (MAF) less than 0.01% were subsequently removed. Finally, 9,814,944 loci remained in the dataset.

## Methods

### Inclusion and ethics declarations

The TWB obtained written informed consent from all participants. The TWB (TWBR10911-01 and TWBR11005-04) and the Institute Review Board at Academia Sinica (AS-IRB01-17049 and AS-IRB01-21009) approved our data application and use.

### Image quality control and pre-processing

#### ABD images

For ABD medical image quality control and pre-processing, we employed the following steps for image quality control sequentially (**Fig. S2A**): (1) Removal of images with inconsistent sizes compared to others (m = 522 images); (2) Exclusion of images without pixel content (m = 127 images); (3) Elimination of B-mode images with annotations (m = 65,718 images); (4) Removal of Doppler mode images (m = 3,829 images); (5) Removal of entirely black images (m = 2 images); (6) Removal of images with multiple windows (m = 2,090 images); (7) Exclusion of images that are not ABD (m = 52 images). After applying these exclusion criteria, it remains m = 547,162 images from n = 22,062 participants. Subsequently, we employed the following step for image pre-processing sequentially (**Fig. S2A**): (1) Conversion of RGB images into grayscale; (2) Cropping and selection of the Region of Interest (ROI) and exclusion of the surrounding text. After cropping the ABD medical images, the resulting image size was shortened from 614 x 816 pixels to 496 x 685 pixels (width x height) (**Fig. S2A**).

#### BMD images

For BMD medical image quality control and pre-processing, we employed the following steps for image quality control procedures sequentially (**Fig. S2B**): (1) Cropping and selecting the Region of Interest (ROI) and excluding the surrounding text. (2) Remove the existing contours or bounding box from the images (m = 22,071). After applying these exclusion criteria, m = 21,725 spine images, 21,749 left hip images, and 21,749 right hip images remained. For spine images, we removed images with fewer than three vertebrae or image size height <163 pixels (**Fig. S2B**). Finally, the detail of BMD image size after pre-processing was shown, and a padding method was used to ensure consistent image sizes across different images (**Supplementary Text 1**).

### Image activation vector extraction and classification for T2D

Convolution-based DenseNet121 algorithm ^30^ was applied for activation vector extraction based on the first training data (i.e., **Dataset 1** in **Fig. 1B**) and validation data (i.e., **Dataset 3** in **Fig. 1B**). For ABD medical image, Average Activation Vector (AAV) or Graph Neural Networks (GNN) was applied for integrative activation vectors from the same sample’s several ABD medical images. XGBoost algorithm ^28^ was applied to classify T2D based on imaging activation vectors, genetic PRS, and demographic variables. Classification was trained and validated based on the second training data (i.e., **Dataset 2** in **Fig. 1B**) and validation data (i.e., **Dataset 3** in **Fig. 1B**). All classification models were tested based on the first testing data (i.e., **Dataset 4** in **Fig. 1B**). The best model was further validated based on the second independent testing dataset (i.e., **Dataset 5** in **Fig. 1B**).

The DenseNet121 architecture was shown (**Fig. 2A**). The DenseNet121 models were trained with the following settings: image size (64 x 64, 224 x 224, 256 x 256), batch size (64, 128, 256), and pre-trained (w/ or w/o). All the DenseNet121 models were trained using an initial learning rate of 0.05 for 500 epochs. The best parameters set was used.

**Figure 2.**
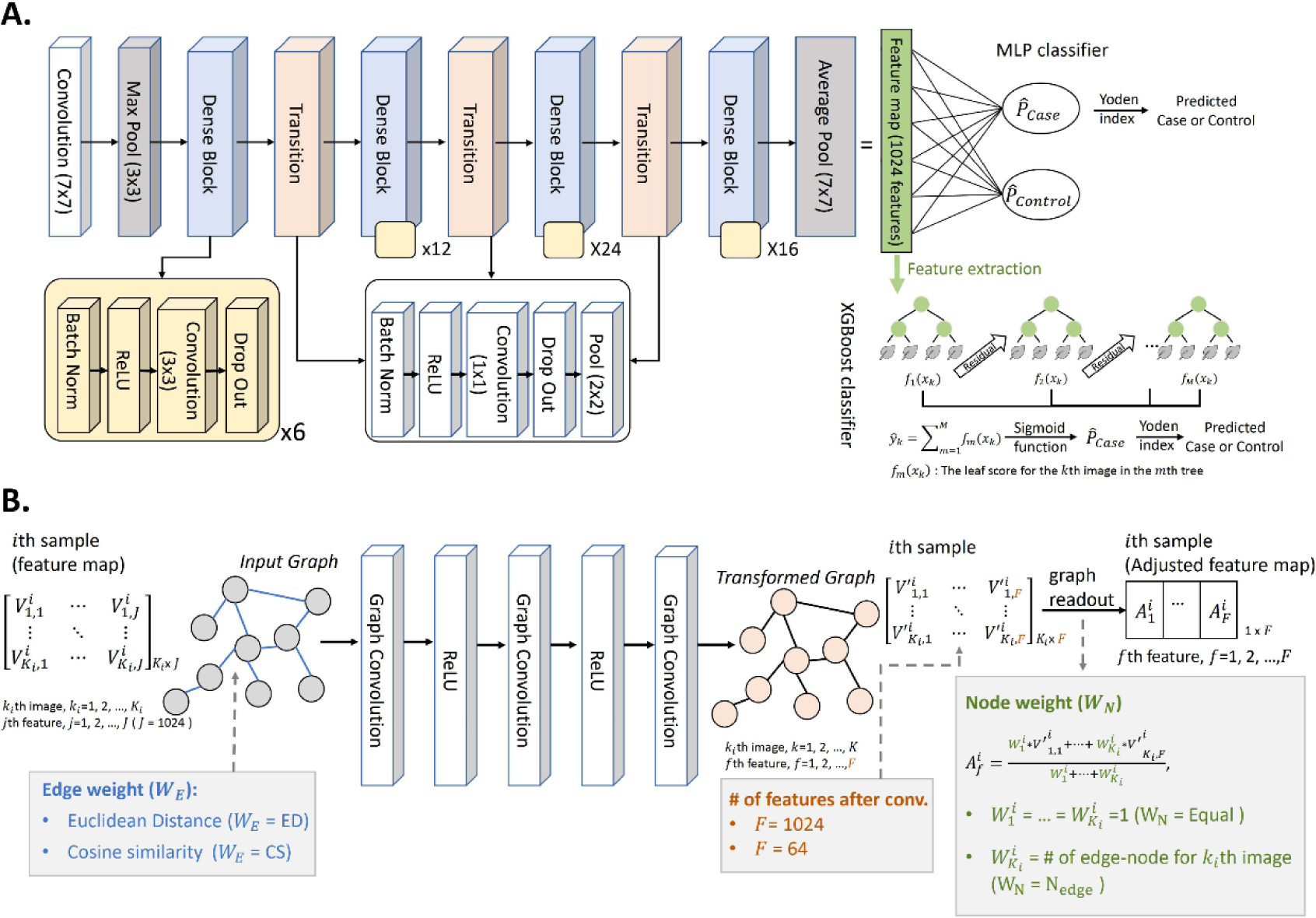
Architecture diagrams. **(A) Densely Connected Convolutional Neural Networks with 121 layers (DenseNet-121). (B) Graph Neural Network (GNN).**

The GNN architecture was shown (**Fig. 2B**). For GNN, we obtained localized node embedding using “GNNConv” ^31^ applied from PyTorch Geometric (PyG) ^32^ and used mean or weighted mean pooling for a graph readout before applying classifier. The GNN models were trained with the following parameters: edge weight calculated by Euclidean distance or cosine similarity, number of features after convolution (64 or 1024), and graph readout according to node weight given equal weight (mean pooling) or weighted by number of edge node. The best distance cutoff between two nodes determines whether an edge exists between two nodes. All the GNN models are trained using an initial learning rate 0.05 for 500 epochs. The best parameters set was used.

The XGBoost models ^28^ were trained with the following default parameter settings: maximum depth equal to 6, learning rate equal to 0.3, the value of the regularization parameter alpha (L1) was set to 0, and lambda (L2) was set as 1, the number of boosting stages was 100, and the early-stop parameter was set to 30. Parameter tuning was conducted to establish the best model (**Supplemental Table S2**).

The model’s effectiveness was evaluated by computing the area under the receiver operating curve (AUC). The performance of the created models was assessed using accuracy, sensitivity, specificity, and Youden index metrics. The optimal threshold value for the XGBoost model on the validation data was determined using the Youden index ^33^.

### Multi-image Risk Score and Polygenic Risk Score

#### Multi-image Risk Score (MRS)

The computation of the MRS involved a sophisticated process. Initially, imaging pixels were utilized as input, and an activation vector was meticulously derived and consolidated through a series of operations, including convolution, pooling, transition, and dense block within the DenseNet121 architecture (refer to **Fig. 2A** for an illustration). Subsequently, this activation vector, also known as the feature vector, was constructed. The feature vector was the input variable for assessing the T2D disease status employing the XGBoost classifier^28^. Notably, the XGBoost feature importance algorithm was applied to identify crucial features, and the MRS was ultimately calculated as the likelihood of an individual being classified as a T2D case.

#### Polygenic risk score (PRS)

The PRS construction closely followed the methodology in our prior investigation ^25^. PRS-CSx ^34^ was employed, utilizing meta-GWAS summary statistics for T2D across diverse ancestral populations. Specifically, the data encompassed East Asian (EAS) populations (56,268 cases and 227,155 controls from the DIAGRAM Consortium ^35^), European (EUR) populations (80,154 cases and 853,816 controls from the DIAGRAM Consortium ^35^), and South Asian (SAS) populations (16,540 cases and 32,952 controls from the DIAGRAM Consortium ^35^). Additionally, Linkage Disequilibrium (LD) references from the 1000 Genomes Project ^36^ for each of the three populations (EAS, EUR, and SAS) were incorporated. Weights of 884,327, 880,098, and 900,047 SNPs for EAS, EUR, and SAS were applied to our genotype data to calculate the population-specific PRS using the PLINK (--score command) tool. Subsequently, we combined the population-specific PRS with equal weights to derive the final PRS. The R language was utilized to standardize the PRS, setting the mean to 0 and the standard deviation to 1.

## Results

### All T2D risk evaluation models

We constructed 12 T2D risk evaluation models (***M*1**–***M*12** in **Table 1**) by various combinations of conditions, including imaging type (ABD and BMD), image analysis unit (sample-based and image-based), image analysis type (FECA vs. PIXA), and analysis modality (MUMA vs. SIMA).

**Table 1.** Models and performance.

| No. | Model description | Image type | Image<br>analysis unit | Image data<br>analysis type | Analysis<br>modality | AUC | Accuracy | Sensitivity | Specificity | Youden |
| --- | --- | --- | --- | --- | --- | --- | --- | --- | --- | --- |
| **M1** | $ABD^{IDF}$ | ABD | sample-based | FECA | SIMA | 0.719 | 0.703 | 0.643 | 0.714 | 0.357 |
| **M2** | $ABD_{AVE}^{PIX}$ | ABD | sample-based | PIXA | SIMA | 0.881 | 0.847 | 0.746 | 0.864 | 0.610 |
| **M3** | $ABD_{GNN}^{PIX}$ | ABD | sample-based | PIXA | SIMA | 0.887 | 0.842 | 0.765 | 0.855 | 0.620 |
| **M4** | $ABD_{IMAGE}^{PIX}$ | ABD | image-based | PIXA | SIMA | 0.808 | 0.735 | 0.728 | 0.736 | 0.464 |
| **M5** | $BMD^{IDF}$ | BMD | sample-based | FECA | SIMA | 0.847 | 0.763 | 0.783 | 0.759 | 0.542 |
| **M6** | $BMD_{COMBINE}^{PIX}$ | BMD | sample-based | PIXA | MUMA | 0.824 | 0.774 | 0.697 | 0.788 | 0.485 |
| **M7** | $BMD_{SPINE}^{PIX}$ | BMD | sample-based | PIXA | SIMA | 0.765 | 0.725 | 0.653 | 0.737 | 0.390 |
| **M8** | $BMD_{L.HIP}^{PIX}$ | BMD | sample-based | PIXA | SIMA | 0.763 | 0.736 | 0.649 | 0.751 | 0.400 |
| **M9** | $BMD_{R.HIP}^{PIX}$ | BMD | sample-based | PIXA | SIMA | 0.757 | 0.786 | 0.528 | 0.828 | 0.356 |
| **M10** | $ABD^{IDF} + BMD^{IDF}$ | ABD+BMD | sample-based | FECA | MUMA | 0.871 | 0.830 | 0.745 | 0.845 | 0.590 |
| **M11** | $ABD_{GNN}^{PIX} + BMD_{COMBINE}^{PIX}$ | ABD+BMD | sample-based | PIXA | MUMA | 0.904 | 0.880 | 0.706 | 0.910 | 0.616 |
| **M12** | $ABD_{GNN}^{PIX} + BMD_{COMBINE}^{PIX} + G + D$ | ABD+BMD | sample-based | PIXA | MUMA | 0.944 | 0.868 | 0.889 | 0.865 | 0.754 |

Abbreviation list. ABD: abdominal ultrasonography; BMD: bone density scan; FECA: feature-centric analysis; PIXA: pixel-based analysis; SIMA: single-modality analysis;
MUMA: multi-modality analysis; G: genetic predictor (PRS); D: demographic variables (age, sex, and family history of T2D).

### Pixel-based analysis (PIXA) demonstrated competitiveness compared to feature-centric analysis (FECA)

We constructed T2D risk evaluation models utilizing raw pixel or IDF data of ABD and BMD medical imaging as predictors. In the case of ABD, compared to FECA (Model ***M*1** in **Table 1**), PIXA (Model ***M*2** in **Table 1**) outperformed across all performance metrics (**Table 1** and **Fig. 3A**). In the case of BMD, compared to the FECA (Model ***M*5** in **Table 1**), PIXA (Model ***M*6** in **Table 1**) demonstrated similar performance (see **Table 1** and **Fig. 3B**). The results highlight that PIXA offered valuable information for T2D risk evaluation, even in the absence of consideration of clinical data from medical technologists.

**Figure 3.**
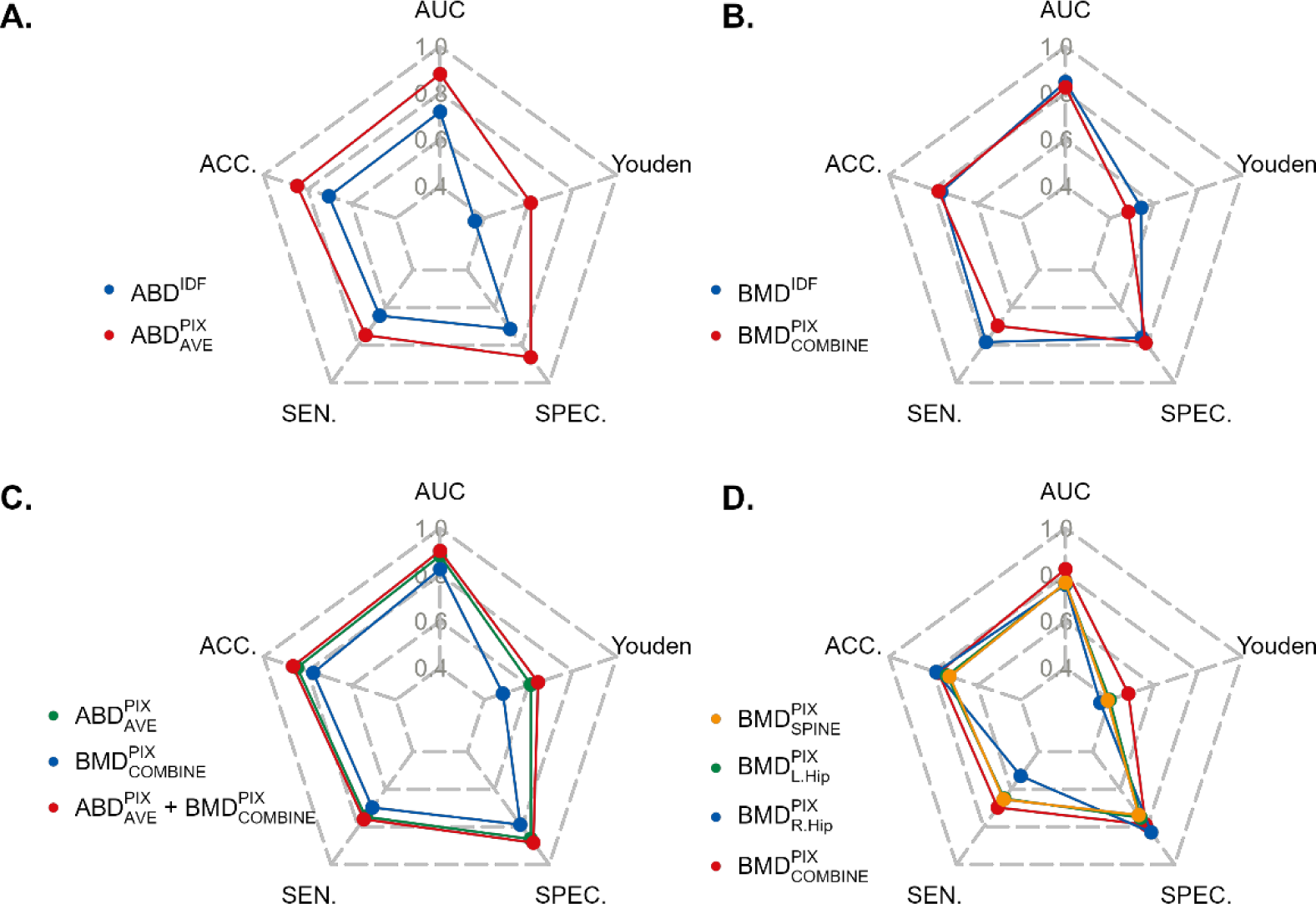
Model Comparison. **(A) Comparison of PIXA** 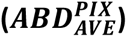 **and FECA (*ABD^IDF^*) in ABD imaging analysis.** PIXA exhibited superior performance in T2D risk evaluation compared to FEXA. **(B) Comparison of PIXA** 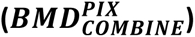 **and FECA (*BMD^IDF^*) in BMD imaging analysis.** PIXA and FECA exhibited similar performance in T2D risk evaluation. **(C) Comparison of multi-modality analysis (MUMA) and single-modality analysis (MUMA) in ABD and BMD imaging analysis.** MUMA of ABD and BMD outperforms SIMA of ABD and SIMA of BMD. **(D) Comparison of multi-modality analysis (MUMA) and single-modality analysis (MUMA) in different BMD images, including spine, left hip, and right hip.** MUMA of the three types of BMD images outperforms SIMA of each of the three types of BMD images.

### Multi-modality analysis (MUMA) outperformed single-modality analysis (SIMA)

#### Combining ABD and BMD imaging

A multi-modality PIXA, which concurrently analyzed the raw pixel data of both ABD and BMD imaging data (Model ***M*11** in **Table 1**), consistently outperformed the individual-modality PIXA of ABD imaging (Model ***M*3** in **Table 1**) and BMD imaging (Model ***M*6** in **Table 1**) across majority of performance measures, particularly in AUC, ACC, SPEC, and Youden index (refer to **Fig. 3C**). The results underscore the enhanced performance of a MUMA compared to a SIMA, although caution is advised due to the associated higher data-collection cost in a MUMA. A similar finding applies to FECA: multi-modality FECA (Model ***M*10** in **Table 1**) outperforms a single-modality FECA: ABD imaging (Model ***M*1** in **Table 1**) and BMD imaging (Model ***M*5** in **Table 1**) (**Fig. S3**).

#### Combining spine, left hip, and right hip BMD imaging

As to BMD imaging, which consists of the spine (Model ***M*7** in **Table 1**), left hip (Model ***M*8** in **Table 1**), and right hip (Model ***M*9** in **Table 1**) medical imaging, the three individual PIXAs exhibited a close performance in T2D classification (**Fig. 3D**). A MUMA which integrated spine, left hip, and right hip medical imaging (Model ***M*6** in **Table 1**) exhibited an improvement compared to the three individual SIMAs (Models ***M*7–*M*9** in **Table 1**), particularly in increased AUC, SEN, and Youden index (**Fig. 3D**). The results once again illustrated a better performance for a MUMA compared to a SIMA.

### Robustness analysis

#### Robustness analysis of DenseNet121 parameters suggests the constructed models are robust

Our robustness analysis for ABD imaging examined three image sizes (64×64, 128×128, and 224×224), three batch sizes (64, 128, and 256), and usage of the pre-trained model (yes or no) (**Fig. 4A**). All combinations of these settings demonstrated similar performance in terms of AUC. The model attaining the highest AUC of 0.800 is characterized by an image size of 224 x 224 pixels, batch size of 64, and no pre-trained weights (**Fig. 4A**). These parameters were fixed in our subsequent analysis. Other parameters in DenseNet121 were set as default values. The final parameters were also applied to BMD imaging.

**Figure 4.**
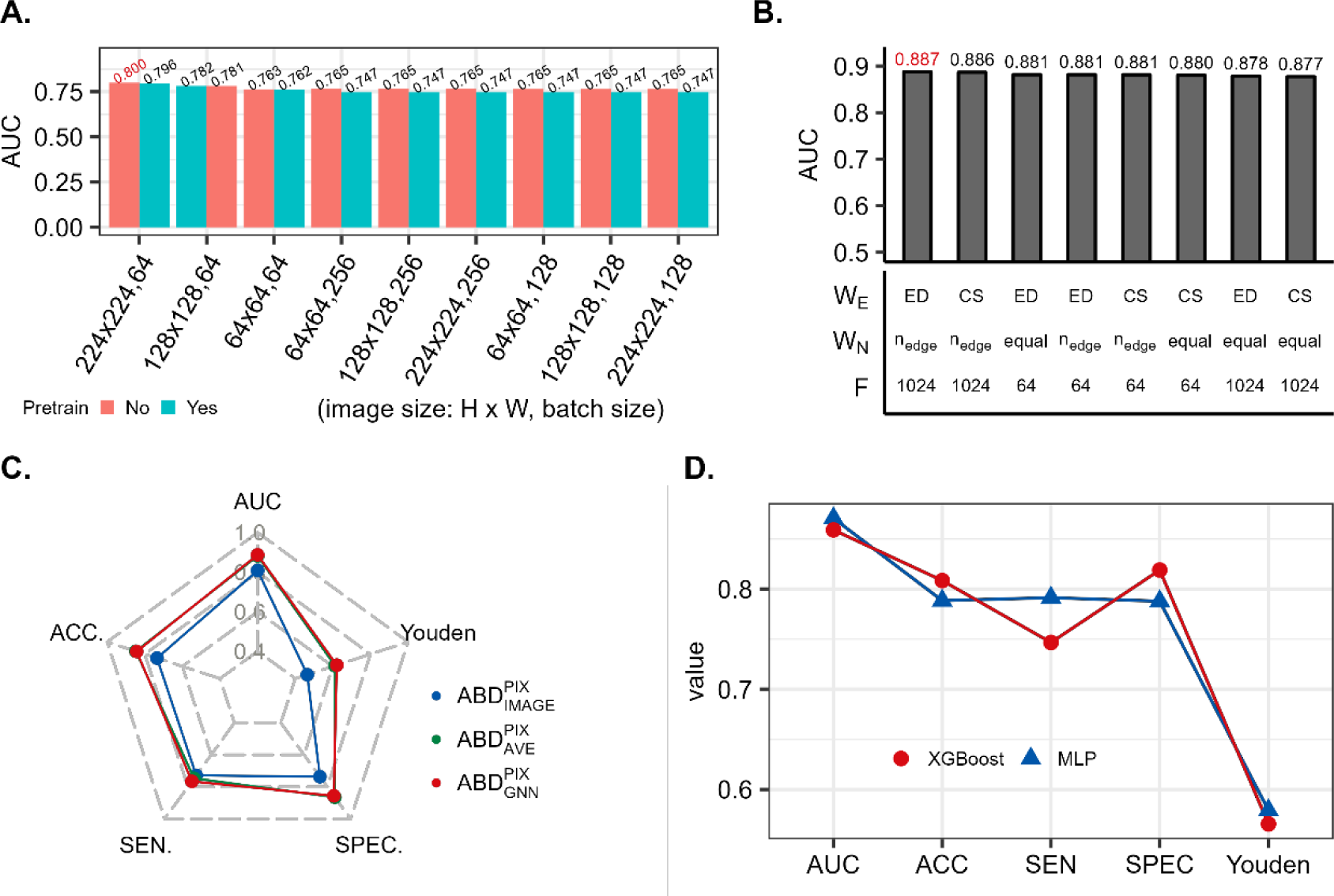
Results of robustness analysis in ABD imaging. **(A) The different parameter settings of the DenseNet121 model for the ABD imaging analysis.** Testing AUC results indicate that the configuration with an image size of 224 x 224, a batch size 64, and a pre-trained model set to False performed the best. (B) The different parameter settings of the GNN model for the ABD imaging analysis. *W_eg_* refers to the weight of the edge used (Euclidean Distance - ED or Cosine Similarity - CS). *W_n_* represents the weight of nodes, either equal weight or weighted by the number of edge nodes (*n_edge_*). *F* indicates the number of features after graph convolution. The testing AUC shows that the best performance is achieved with settings: *W_eg_* = ED, *F* = 1024, and *W_n_* = *n_edge_*. (C) Comparative analysis between image-based analysis, sample-based analysis with average weights, and sample-based analysis with GNN weights in ABD imaging analysis. The sample-based analysis with GNN weights performs best. **(D) Comparison of two classifiers, Multilayer Perceptron (MLP) and eXtreme Gradient Boosting (XGBoost).** MLP and XGBoost exhibited similar performances in all measures: AUC, ACC, SEN, SPEC, and Youden index.

#### Robustness analysis of Graph Neural Network parameters suggests the constructed models are robust

GNN was applied to account for the intra-individual dependency of multiple images of ABD available for each individual. Our robustness analysis for GNN considered two edge weights *W_E_* (“Euclidean distance” and “Cosine similarity”), two node weights *W_N_* (“Equal-weight” and “Number of edges connected to a node”), and two numbers of features after the graphical convolution (64 or 1,024) (**Fig. 4B**). All combinations of these settings exhibited similar performance in AUC, suggesting that the constructed GNN models are robust. The model attaining the highest AUC of 0.887 is characterized by Euclidean-distance edge weight (*W_E_* = ED), 1,024 features after convolution (*F* = 1,204), and node weight proportional to the number of edge nodes (*W_N_* = *n*_edge_) (**Fig. 4B**). Existence of an edge/link between two nodes was determined by a threshold “ED”. The results show that the optimal ED cutoff, which attained the highest AUC of 0.887, was ED = 0.5 (**Fig. S4**).

#### Robustness analysis suggests the sample-based method outperforms the image-based method and accounting for within-sample correlation further improves performance

The results suggest that the sample-based method (Models ***M***2 and ***M***3 in **Table 1**), which integrates multiple images of each individual, outperforms the image-based method (Model ***M***4 in **Table 1**) (**Fig. 4C**). Furthermore, a sample-based method using GNN (i.e., the model with accounting for within-sample image correlation) (Model ***M***3 in **Table 1**) is slightly better than the sample-based method using a direct cross-image average (i.e., without accounting for image correlation) (Model ***M***2 in **Table 1**) (**Fig. 4C**).

#### Robustness analysis of classifiers (XGBoost and MLP) have similar performance, suggesting the finding is robust

To examine the robustness of our findings concerning classifiers, in addition to the eXtreme Gradient Boosting (XGBoost) classifier, we also compared it with the multi-layer perception (MLP) classifier based on the ABD imaging. The results show that XGBoost and MLP exhibited similar results in terms of AUC, ACC, SEN, SPEC, and Youden index (**Fig. 4D**), which were averaged across three PIXA variants – sample-based PIXA with an average weight (Model ***M*2** in **Table 1**), sample-based PIXA with a GNN weight (Model ***M*3** in **Table 1**), and image-based PIXA (Model ***M*4** in **Table 1**). The detailed results of the three models based on ABD (**Fig. S5A**) and BMD (**Fig. S5B**) are demonstrated.

### Clinical consideration

#### Integrative model

In addition to the imaging data, other crucial features for T2D diagnosis including genetic component (PRS) and demographic variables (sex, age, and family history of T2D) were also considered in the ABD genetic–imaging integrative analysis (**Fig. 5A-1**) and BMD genetic–imaging integrative analysis (**Fig. 5A-2**), and ABD+BMD integrative analysis (**Fig. 5A-3**); additional results for the spine, left hip, and right hip are presented (**Figs. S6A-1 – S6A-3**). Considering five performance metrics, the combination of imaging features, demographic factors, and genetic PRS performed the best. Imaging features performed exceptionally well, surpassing the performance of demographic characteristics and genetic PRS when considered individually, particularly in the ABD analysis.

**Figure 5.**
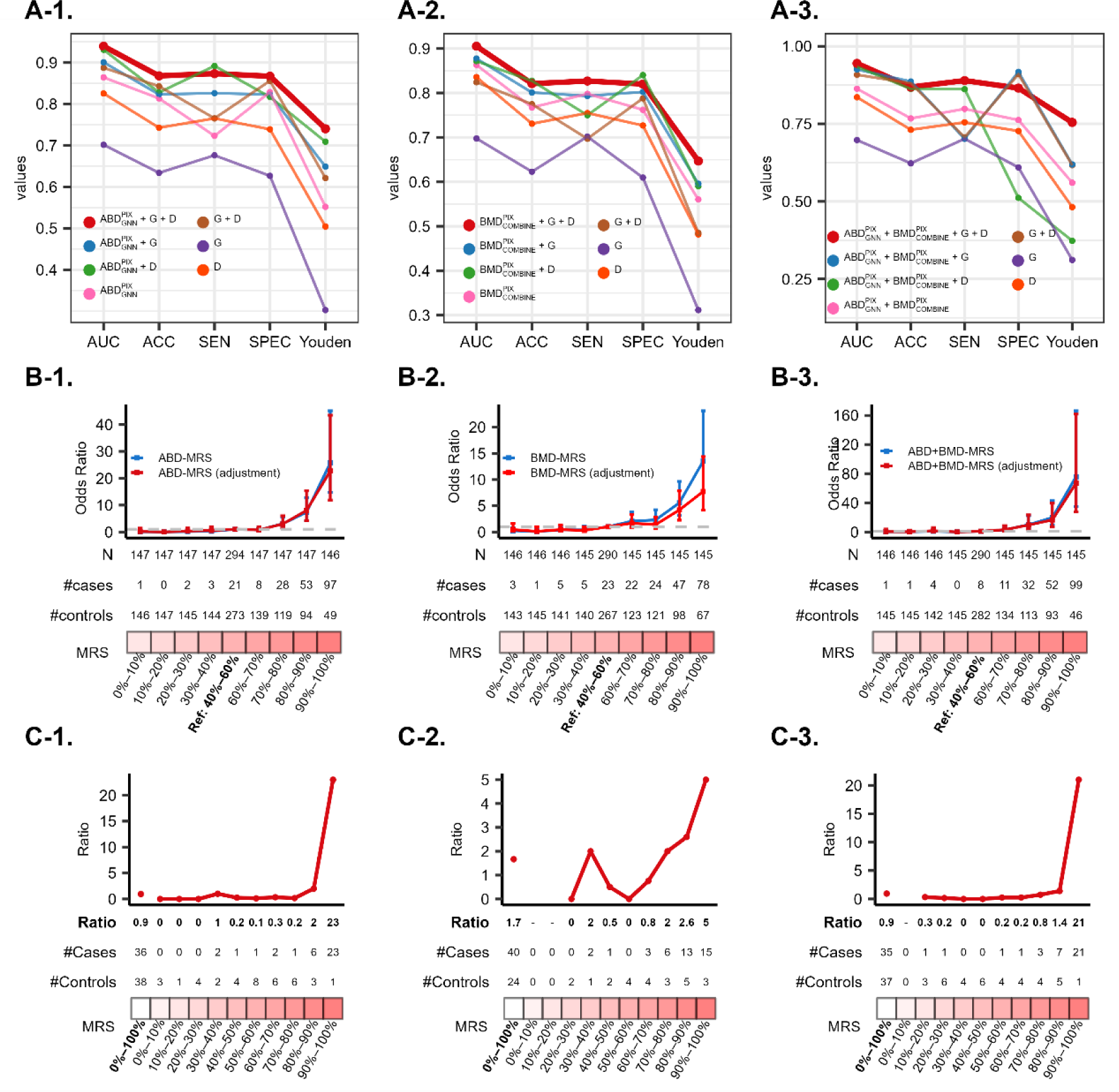
Risk evaluation models for T2D. **(A) Performance of various models accounting for genetic PRS (G), demographic variables (D) – age, sex, and T2D family history (D), and medical images of ABD (A-1), BMD (A-2), and ABD+BMD (A-3).** The model comprising G, D, and medical images performs the best in T2D risk evaluation. **(B) Positive correlation between MRS and T2D odds ratio for ABD (B-1), BMD (B-2), and ABD+BMD (B-3).** In each decile of MRS based on the image, the odds ratio of T2D risk and its 95% confidence interval were calculated based on an unadjusted model (blue line) and model adjusted by age, sex, and T2D family history (red line), where the MRS group in 40%–60% is set as the reference group. **(C) Identification of high-risk subgroup based on MRS of ABD (C-1), BMD (C-2) and ABD+BMD (C-3).** For ABD imaging and ABD+BMD imaging, the high-risk group was females older than 62 with a T2D family history, and their MRS group was 90%– 100%. For BMD imaging, in addition to being identical to the group identified by ABD-based MRS, another high-risk group was men older than 62 with a family history of T2D.

#### Best risk assessment model for T2D

The best medical imaging model (***M*11** in **Table 1**) is a sample-based PIXA and MUMA, which combines ABD and BMD imaging (spine, left hip, and right hip imaging). On top of the medical imaging in this model, demographic variables (D) and genetic components (G) were further included. Finally, the best risk evaluation model is 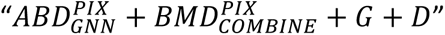 (***M*12** in **Table 1**), attaining AUC = 0.944, ACC = 0.868, SEN = 0.889, SPE = 0.865, and Youden index = 0.754 in the first testing data based on a classification threshold 0.01. The model was further validated well in the second independent testing dataset with AUC = 0.954, ACC = 0.875, SEN = 0.882, SPE = 0.875, and Youden index = 0.757. The XGBoost model parameter tuning and performance evaluation for this model are shown (**Supplementary Table S2**).

#### Multi-image risk score (MRS)

ABD-based and BMD-based multi-image risk scores (MRSs) were calculated for each participant. The odds ratio of T2D and its corresponding confidence interval revealed a positive correlation between MRS and T2D (**Fig. 5B-1** for ABD and **Fig. 5B-2** for BMD). This suggests that the risk of developing T2D increases with higher MRS values than the reference group: 40%–60% decile group of MRS. These findings could potentially lead to more effective treatments in the future. Similar results can also be found at the SIMA of the spine (**Fig. S6B-1**), left hip (**Fig. S6B-2**), and right hip (**Fig. S6B-3**).

#### Identification of high-risk subgroups using MRSs

Furthermore, we identified specific high-risk subgroups within the study population. In the analyses of ABD, BMD, and ABD+BMD, consistently high T2D risk (i.e., the ratio of the number of cases vs. controls) was observed within the high MRS group (90– 100% decile group) for both women and men aged older than 62 years with a family history of T2D, as follows: In the ABD analysis, T2D risks were 23 in the female group (**Fig. 5C-1**) and 8 in the male group. In the BMD analysis, T2D risks were 5 in the male group (**Fig. 5C-2**) and 4.4 in the female group. In the ABD+BMD analysis, T2D risks were 21 in the female group (**Fig. 5C-3**) and 8 in the male group. These T2D risks in the 90–100% MRS decile group were significantly higher than those in the lower-MRS groups.

## Discussion

### High-performance genetic-imaging integrated analysis of T2D risk evaluation and diagnosis

In our previous work ^25^, which considered an AI-enhanced integration of genetic and medical imaging data for T2D risk assessment, a FECA analysis based on the IDFs of four medical images (ABD, BMD, CAU, and ECG) in the Taiwan Biobank achieved an AUC of 0.880, increasing to 0.945 after incorporating demographic (age and family history of T2D) and genetic information (PRS). The current study, focusing on two T2D-related medical images (ABD and BMD), demonstrates that PIXA outperforms FECA for T2D risk evaluation. Despite analyzing fewer types of images than our previous study ^25^, this investigation achieved an AUC of 0.902 based on ABD and BMD pixel data. The AUC further increased to 0.953 after incorporating demographic and genetic information. The result underscores the potential of an integrated multi-modality study of genetic analysis and medical imaging PIXA for precision medicine.

For precision medicine, genetic information (genome-wide SNPs) and medical imaging data (image-wide pixels) provide individualized information compared to traditional biochemistry and body measurement indices, such as HbA1c and fasting glucose have demonstrated limitations in disease risk prediction, as highlighted in various studies ^37, 38, 39, 40, 41, 42, 43, 44^. These metrics often show limited sensitivity, especially in specific populations, and lack accuracy in predicting pre-diabetes and T2D. Genetic data offers stability for evaluating disease risk and diagnosis in precision medicine, while medical imaging data provides detailed multi-modality information for human organs, contributing to stable and insightful disease risk evaluation, diagnosis, and classification.

### Comparison of pixel-based analysis (PIXA) and feature-centric analysis (FECA)

Our investigation reveals that PIXA demonstrates competitive and, in some cases, superior performance compared to FECA in T2D classification (**Figs. 3A** and **3B**). In the specific case examined in this paper, two potential explanations warrant careful consideration. Firstly, it is plausible that IDFs did not entirely extract the complete information embedded within the raw pixel data. Specifically, only 28 IDFs are extracted in the case of ABD, and some of these features may lack direct association with T2D. Secondly, IDFs defined by medical technologists might be suboptimal. For instance, accurately labeling the exact fatty liver level (normal, mild, moderate, and severe) poses challenges, particularly for closely related levels at the borderline. Consequently, PIXA, without labor-intensive labeling and expensive annotation, offers precise feature quantification with artificial intelligence, providing a high-performance solution for disease classification and risk assessment. This approach paves the way for effective and practical clinical applications.

### Comparison of MUMA and SIMA

Our investigation reveals that a MUMA provides superior performance compared to a SIMA (**Figs. 3C and 3D**) if there is non-overlapping information in the images of different modalities. Our integrated analysis of ABD and BMD medical imaging outperforms individual analyses of ABD and BMD (**Fig. 3C**). The enhanced performance in the integrated analysis can be attributed to the non-overlapping contributions of ABD and BMD to T2D. Conversely, our integrated classification analysis of the spine, left hip, and right hip medical imaging performed similarly to the three individual analyses (**Fig. 3D**). This suggests that these imaging modalities provide highly correlated and redundant information for T2D despite representing different body sections. These observations underscore the importance of carefully selecting and integrating imaging modalities for disease classification and considering each modality’s unique contributions to enhance overall diagnostic accuracy. Our findings align with previous studies demonstrating the superiority of MUMA over SIMA in disease classification ^45, 46, 47, 48^.

### Robustness analysis for FECA

We conducted a robustness analysis for the FECA, considering various factors such as pre-trained models, image sizes, batch sizes, and classifiers. Firstly, employing a pre-trained DenseNet121 model ^49, 50, 51, 52^ did not enhance performance (**Fig. 4A**), potentially due to differences in characteristics between ImageNet ^53^ and medical imaging data and the limited number of images in ImageNet. Secondly, variations in image sizes and batch sizes demonstrated minimal impact on AUCs (**Fig. 4A**). Thirdly, variations in edge weight methods – “Euclidean distance (DS)” and “Cosine similarity (CS),” node weight methods – “Equal weight (EW)” and “Unequal weight proportional to the number of nodes connected to”), and several nodes after convolution exhibited close AUCs (**Fig. 4B**). Thirdly, GNN which considers within-individual image correlation performed slightly better than AVE which does not consider the correlation (**Fig. 4C**); however, the difference in performance is limited. Finally, the alternative classifier – multilayer perceptron (MLP), exhibited no significant difference in performance compared to XGBoost (**Fig. 4D**). This consistency across different classifiers underscores the robustness of our findings, enhancing the credibility and generalizability of our proposed approach for T2D risk evaluation and classification.

## Conclusion

In conclusion, this study highlights the compelling findings that applying artificial intelligence, comprising deep learning and machine learning, to integrated genetic and medical imaging PIXA provides a fully automated, low-labor, cost-saving, and high-accuracy analysis. Incorporating multi-modality data, encompassing diverse-dimensional information, significantly enhances the performance compared to single-modality data analysis. Notably, medical imaging PIXA emerges as a competitive and, in many instances, superior performer compared to FECA. Integrating genome-wide genetic data with multi-modality imaging marks a revolutionary advancement in precision medicine and smart health for T2D. These results provide crucial insights into the potential transformative impact of advanced analytical methodologies on the future of T2D diagnosis and personalized healthcare.

## Data Availability

The data analyzed in this study were obtained from the Taiwan Biobank with proper approval. The Taiwan Biobank retains ownership rights, so the data have not been deposited in a public repository. Researchers interested in accessing the data must apply through the Taiwan Biobank's formal process. Detailed instructions for data access requests can be found on the Taiwan Biobank's official website (https://www.twbiobank.org.tw/index.php). This paper provides Source data in the Supplementary Information and Source Data files. Meta-GWAS summary statistics of T2D in multiple populations from the DIAGRAM Consortium are available at https://diagram-consortium.org/downloads.html. The linkage disequilibrium reference from various populations of the 1000 Genomes Project can be downloaded from https://github.com/getian107/PRScsx.

https://diagram-consortium.org/downloads.html

https://github.com/getian107/PRScsx

## Acknowledgments

This work was supported by research grants from the Academia Sinica (AS-PH-109-01 and AS-SH-112-01). Data application and use were approved by the Taiwan Biobank and the Institute Review Board (AS-IRB01-17049 and AS-IRB01-21009). We gratefully acknowledge the Taiwan Biobank for providing the data used in this research. We also extend our thanks to all the participants of the Taiwan Biobank for their invaluable contributions. Technical support in genotyping from the National Center for Genome Medicine of Taiwan is also acknowledged. We thank team members Miss Chih-Ting Yang and Mr. Po-Wen Chen for imaging preprocessing of ABD and BMD and Mr. Chia-Wei Chen and Dr. Shih-Kai Chu for genetic data quality control.

## Author Contributions Statement

H.C.Y. conceptualized and supervised the study. Y.J.H. curated the data and applied software. Y.J.H. & H.C.Y. conducted formal data analysis, visualized the results, and wrote the paper. C.h.C. & H.C.Y. secured funding and provided resources. H.C.Y., Y.J.H., and C.h.C. validated the results.

## Ethics declarations

### Code availability statement

We provide code at the repository at https://github.com/yjhuang1119/Medical_Image_Risk_Assessment_Model for medical image classification and risk assessment using a combination of Densely Connected Convolutional Networks with 121 layers (DenseNet121) and eXtreme Gradient Boosting (XGBoost). The pipeline includes establishing a disease risk assessment model using DenseNet121, extracting feature maps, constructing a final disease risk assessment model using XGBoost, and performance evaluation. The code also computes performance metrics for model evaluation and feature importance scores for model explainability. A README is provided.

## Supplemental information

Supplemental information is available online.

