## Supplemental files for "AI-driven Integration of Multimodal Imaging Pixel Data and Genome-wide Genotype Data Enhances Precision Health for Type 2 Diabetes: Insights from a Large-scale Biobank Study"

### Supplemental Information

| | | |
|---|---|---|
| Figure S1 | Examples of ABD and BMD images in the TWB dataset................. | 2 |
| Figure S2 | Image pre-processing................................................................ | 3 |
| Figure S3 | Comparison of multi-modality FECA and single-modality FECA of ABD and BMD.................................................................................. | 4 |
| Figure S4 | AUC of the cutoff ED................................................................. | 5 |
| Figure S5 | The performance of MLP and XGBoost classifiers in ABD and BMD analysis.............................................................................. | 6 |
| Figure S6 | Clinical consideration of spine, left hip, right hip, and ABD+BMD medical images.......................................................................... | 7 |
| Table S1 | Demographic characteristics, family history, genetic PRS, and image report variables (i.e., image-derived features; IDFs) of the study population.............................................................................. | 8 |
| Table S2 | Comparison between prediction models using default parameters and tuned parameters............................................... | 12 |
| Supp Text 1 | BMD image pre-processing........................................................ | 13 |

### Supplementary Figures

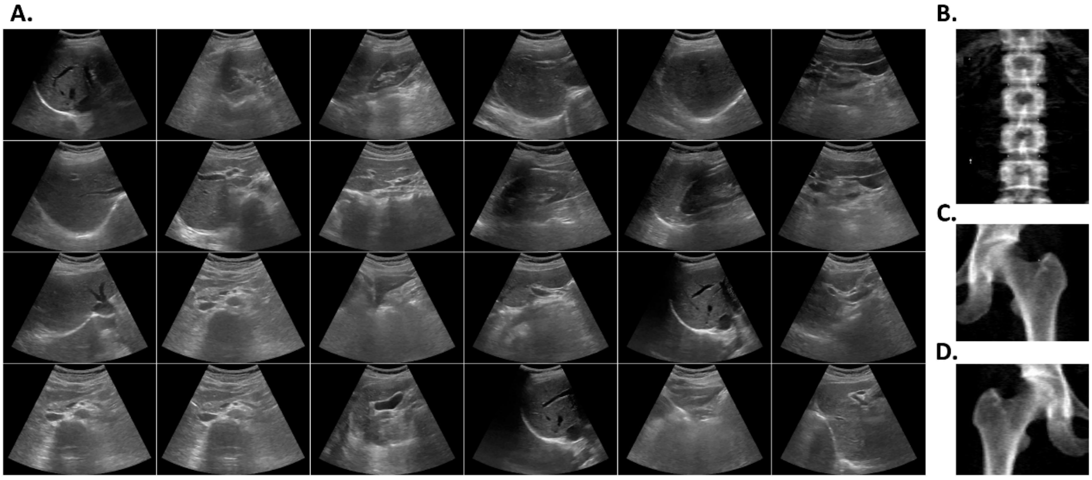


**Figure S1. Examples of ABD and BMD images in the TWB dataset. (A)** The showcased images are sourced from a single participant in the TWB. Each participant possesses different numbers of ABD images, along with one image corresponding to each type of BMD image, including **(B)** spine image, **(C)** left hip image, and **(D)** right hip image.

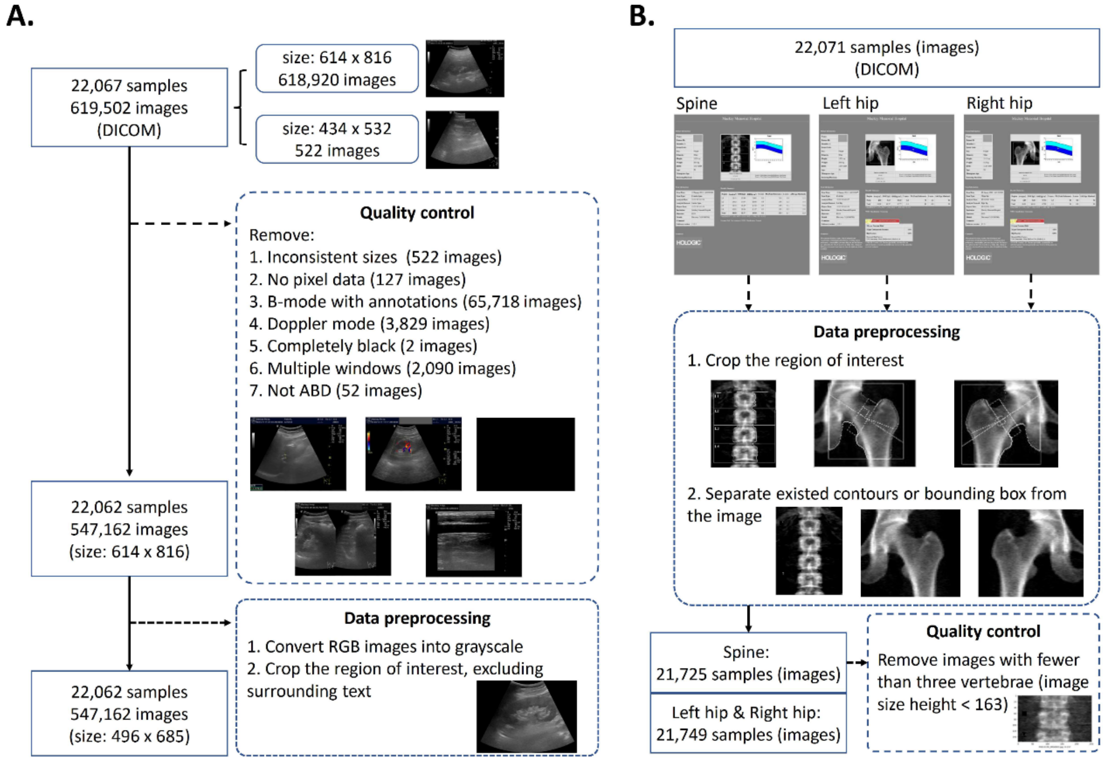


**Figure S2. Image pre-processing. (A) ABD.** Original 22,067 samples with 619,502 images, we employed the following steps for image quality control: Removal of images with inconsistent sizes compared to others (522 images); Exclusion of images with no pixel data available (total: 127 images); Elimination of B-mode images with annotations (total: 65,718 images); Removal of Doppler mode images (total: 3,829 images); Removal of entirely black images (total: 2 images); Removal of images with multiple windows (total: 2,090 images); Identifying and excluding images that are not ABD (total: 52 images); The 22,062 samples with 547,162 images remained. After applying these quality control criteria, we converted RGB images into grayscale and cropped the region of interest while excluding the surrounding text. After cropping the ABD medical images, the resulting image size was 496 x 685 pixels (width x height)**. (B) BMD.** We crop the region of interest for BMD medical images and separate existing contours or bounding boxes from the image. The spine image showing the lumbar region contains five vertebrae; we remove images with fewer than three vertebrae to ensure the quality of the image

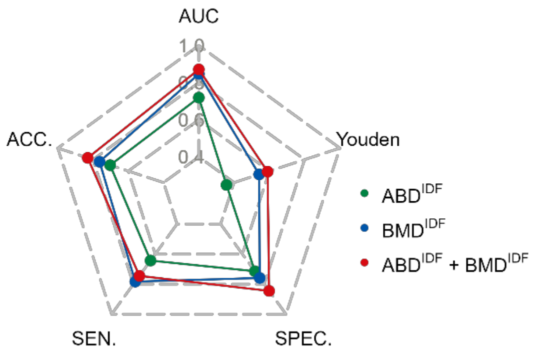


**Figure S3. Comparison of multi-modality FECA and single-modality FECA of ABD and BMD.** Performances of the single-modality FECA of ABD ($\boldsymbol{ABD^{IDF}}$, green), single-modality FECA of BMD ($\boldsymbol{BMD^{IDF}}$, blue), and multi-modality FECA of ABD + BMD ($\boldsymbol{ABD^{IDF} + BMD^{IDF}}$, red). In summary, a multi-modality FECA outperforms a single-modality FECA of ABD and BMD.

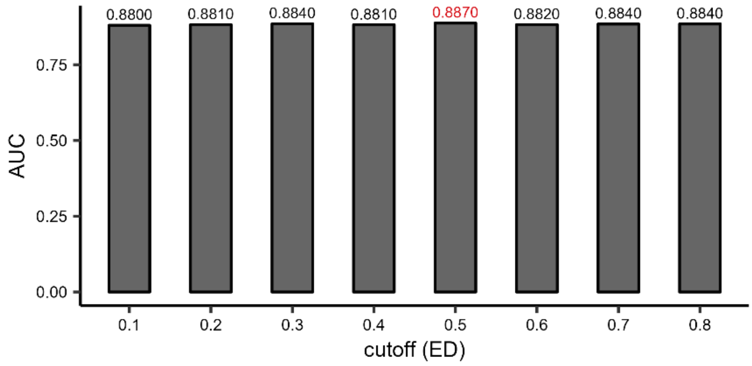


**Figure S4. AUC of the cutoff ED.** The existence of an edge/link between two nodes in the GNN model was determined by a threshold “ED,” a threshold for determining whether an edge between two nodes in the GNN model exists. Thresholds of 0.1–0.8 with an increment of 0.1 were examined. The results show that the optimal ED cutoff, which attained the highest AUC of 0.887, was ED = 0.5.

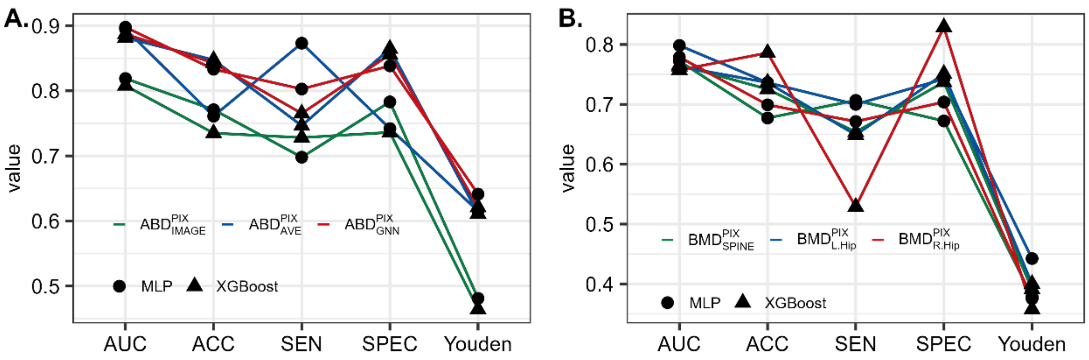


**Figure S5. The performance of MLP and XGBoost classifiers in ABD and BMD analysis. (A) Three ABD prediction methods. (B) Three types of BMD medical imaging.**

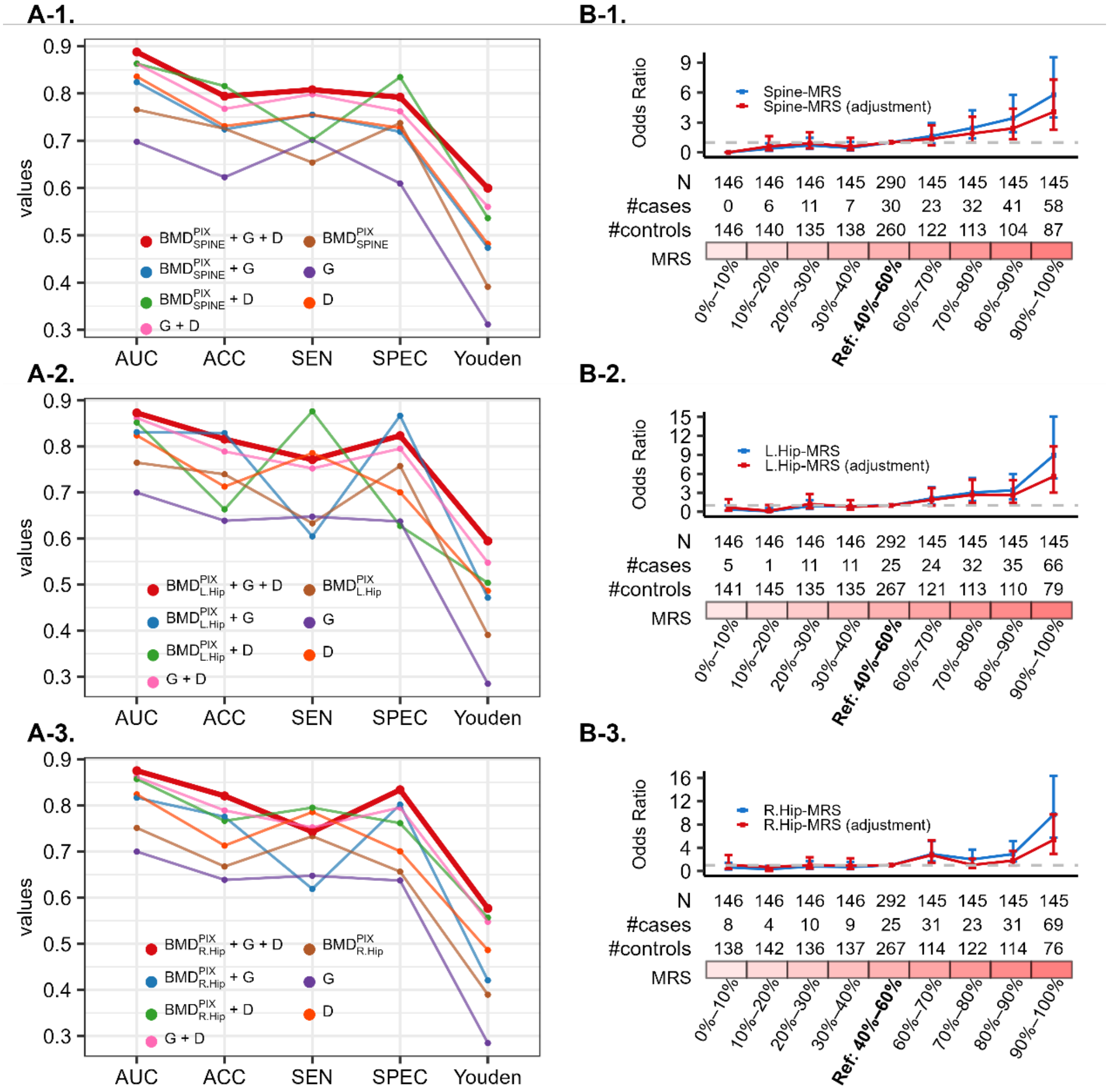


**Figure S6. Clinical consideration of spine, left hip, right hip, and ABD+BMD medical images. (A) Integrative models.** Results of models considering images, genetic PRS (G), and demographic variables, including age, sex, and T2D family history (D), as well as their combinations (A-1. BMD spine; A-2. BMD left hip; A-3 BMD right hip)**. (B) Positive correlation between MRS and T2D odds ratio** (B-1. BMD spine; B-2. BMD left hip; B-3 BMD right hip).

### Supplementary Tables

#### Table S1. Demographic characteristics, family history, genetic PRS, and image report variables (i.e., image-derived features; IDFs) of the study population

| Variable | Overall | T2D group | Non-T2D group | P-value |
|---|---|---|---|---|
| N | 7,342 | 1,066 | 6,276 | |
| **Demographic** | | | | |
| Age, mean (SD) (years) | 53.37 (10.54) | 61.87 (7.56) | 51.93 (10.29) | 1.78E-227 |
| Sex, male, n (%) | 2,590 (35%) | 534 (50%) | 2,056 (32%) | 9.67E-28 |
| Family history | | | | 8.88E-234 |
| 0 | 4,541 (61.8%) | 307 (28.8) | 4,234 (67.5%) | |
| 1 | 2,031 (27.7%) | 382 (35.8) | 1,649 (26.3%) | |
| 2 | 542 (7.4%) | 224 (21.0) | 318 (5.1%) | |
| 3 | 199 (2.7%) | 127 (11.9) | 72 (1.1%) | |
| 4 | 29 (0.4%) | 26 (2.4) | 3 (0.04%) | |
| **Genetic** | | | | |
| Polygenic risk score (PRS) | 0.63 (0.33) | 0.84 (0.31) | 0.59 (0.32) | 1.88E-100 |
| **ABD** | | | | |
| Liver tumor, case, n (%) | 373 (5.1%) | 37 (3.5%) | 336 (5.4%) | 1.20E-02 |
| Liver cyst, case, n (%) | 1,166 (15.9%) | 117 (11.0%) | 1,049 (16.7%) | 2.67E-06 |
| Liver calcification, case, n (%) | 122 (1.7%) | 13 (1.2%) | 109 (1.7%) | 2.75E-01 |
| Liver hemangioma, case, n (%) | 242 (3.3%) | 15 (1.4%) | 227 (3.6%) | 2.69E-04 |
| Liver fibrosis, case, n (%) | 3 (0.0%) | 1 (0.1%) | 2 (0.0%) | 9.16E-01 |
| Chronic liver, case, n (%) | 240 (3.3%) | 36 (3.4%) | 204 (3.3%) | 9.03E-01 |
| Fatty liver, n (%) | | | | |
| Normal | 4,969 (67.7%) | 394 (37.0%) | 4,575 (72.9%) | 3.14E-161 |
| Fatty liver (unknown level) | 1,522 (20.7%) | 313 (29.4%) | 1,209 (19.3%) | |
| Mild | 637 (8.7%) | 271 (25.4%) | 366 (5.8%) | |
| Moderate | 117 (1.6%) | 55 (5.2%) | 62 (1.0%) | |
| severs | 97 (1.3%) | 33 (3.1%) | 64 (1.0%) | |
| Gallbladder polyps, case, n (%) | 635 (8.6%) | 92 (8.6%) | 543 (8.7%) | 1.00E+00 |
| Gallbladder stone, case, n (%) | 478 (6.5%) | 126 (11.8%) | 352 (5.6%) | 4.97E-14 |
| Common bile duct dilatation, case, n (%) | 327 (4.5%) | 66 (6.2%) | 261 (4.2%) | 3.80E-03 |
| Common bile duct stone, case, n (%) | 11 (0.1%) | 1 (0.1%) | 10 (0.2%) | 9.34E-01 |
| Pancreatic cyst, case, n (%) | 11 (0.1%) | 4 (0.4%) | 7 (0.1%) | 1.03E-01 |
| Pancreatic tumor, case, n (%) | 6 (0.1%) | 1 (0.1%) | 5 (0.1%) | 1.00E+00 |
| Pancreatic calcification, case, n (%) | 1 (0.0%) | 0 (0.0%) | 1 (0.0%) | 1.00E+00 |
| Pancreatic calcification, case, n (%) | 3 (0.0%) | 0 (0.0%) | 3 (0.0%) | 1.00E+00 |

| | | | | |
|---|---|---|---|---|
| Pancreatic dilatation, case, n (%) | 14 (0.2%) | 3 (0.3%) | 11 (0.2%) | 7.23E-01 |
| Pancreatitis, case, n (%) | 0 (0.0%) | 0 (0.0%) | 0 (0.0%) | - |
| Spleen tumor, case, n (%) | 4 (0.1%) | 1 (0.1%) | 3 (0.0%) | 1.00E+00 |
| Spleen cyst, case, n (%) | 15 (0.2%) | 3 (0.3%) | 12 (0.2%) | 8.13E-01 |
| Spleen calcification, case, n (%) | 5 (0.1%) | 0 (0.0%) | 5 (0.1%) | 7.74E-01 |
| Spleen accessory, case, n (%) | 7 (0.1%) | 0 (0.0%) | 7 (0.1%) | 5.79E-01 |
| Spleen splenomegaly, case, n (%) | 118 (1.6%) | 33 (3.1%) | 85 (1.4%) | 5.16E-05 |
| Kidney cyst, case, n (%) | 723 (9.8%) | 162 (15.2%) | 561 (8.9%) | 3.28E-10 |
| Kidney stone, case, n (%) | 298 (4.1%) | 72 (6.8%) | 226 (3.6%) | 2.14E-06 |
| Kidney calcification, case, n (%) | 41 (0.6%) | 9 (0.8%) | 32 (0.5%) | 2.58E-01 |
| Kidney hydronephrosis, case, n (%) | 45 (0.6%) | 11 (1.0%) | 34 (0.5%) | 9.23E-02 |
| Kidney polycystic, case, n (%) | 0 (0.0%) | 0 (0.0%) | 0 (0.0%) | - |
| Kidney tumor, case, n (%) | 106 (1.4%) | 13 (1.2%) | 93 (1.5%) | 6.00E-01 |
| **BMD** | | | | |
| Spine k | 1.13 (0.00) | 1.13 (0.00) | 1.13 (0.00) | 8.48E-44 |
| Spine d0 | 46.25 (1.57) | 45.31 (1.65) | 46.41 (1.50) | 5.87E-81 |
| Spine thickness | 6.73 (0.85) | 7.41 (0.93) | 6.62 (0.79) | 4.23E-123 |
| Width of the region of interest in the spine | 116.04 (0.82) | 116.09 (0.47) | 116.04 (0.86) | 5.62E-03 |
| Length of the region of interest in the spine | 133.27 (7.77) | 132.99 (7.95) | 133.31 (7.74) | 2.26E-01 |
| bone area in the lumbar spine (L1) | 12.83 (1.82) | 13.36 (1.97) | 12.74 (1.78) | 6.01E-21 |
| bone mineral content in the lumbar spine (L1) | 11.12 (3.17) | 12.12 (3.75) | 10.95 (3.03) | 2.60E-21 |
| bone mineral density in the lumbar spine (L1) | 0.86 (0.16) | 0.89 (0.17) | 0.85 (0.16) | 2.88E-13 |
| lumbar spine (L1) T-Score | -1.04 (1.39) | -0.86 (1.42) | -1.07 (1.38) | 6.20E-06 |
| lumbar spine (L1) Z-Score | -0.32 (1.28) | 0.15 (1.34) | -0.40 (1.26) | 5.89E-34 |
| The peak reference value for the lumbar spine (L1) | 88.12 (15.82) | 90.22 (16.03) | 87.77 (15.75) | 4.44E-06 |
| Age-matched value for lumbar spine (L1) | 96.07 (16.02) | 102.00 (17.05) | 95.07 (15.62) | 2.10E-33 |
| bone area in the lumbar spine (L2) | 13.80 (1.85) | 14.30 (1.99) | 13.72 (1.82) | 1.54E-18 |
| bone mineral content in the lumbar spine (L2) | 13.00 (3.44) | 13.86 (4.01) | 12.85 (3.32) | 2.51E-14 |
| bone mineral density in the lumbar spine (L2) | 0.93 (0.17) | 0.96 (0.18) | 0.93 (0.17) | 9.35E-06 |
| lumbar spine (L2) T-Score | -1.07 (1.48) | -0.96 (1.50) | -1.09 (1.48) | 7.46E-03 |
| lumbar spine (L2) Z-Score | -0.27 (1.37) | 0.17 (1.40) | -0.34 (1.35) | 2.71E-27 |
| The peak reference value for the lumbar spine (L2) | 88.77 (15.58) | 89.94 (15.68) | 88.57 (15.56) | 8.76E-03 |
| Age-matched value for lumbar spine (L2) | 96.96 (15.78) | 102.04 (16.61) | 96.10 (15.48) | 2.85E-26 |
| bone area in the lumbar spine (L3) | 15.22 (1.90) | 15.61 (2.05) | 15.16 (1.87) | 7.33E-11 |
| bone mineral content in the lumbar spine (L3) | 15.08 (3.71) | 15.82 (4.32) | 14.95 (3.58) | 1.31E-09 |
| bone mineral density in the lumbar spine (L3) | 0.98 (0.17) | 1.00 (0.18) | 0.98 (0.17) | 2.94E-04 |
| lumbar spine (L3) T-Score | -0.98 (1.52) | -0.83 (1.61) | -1.00 (1.50) | 1.80E-03 |
| lumbar spine (L3) Z-Score | -0.15 (1.41) | 0.33 (1.49) | -0.23 (1.39) | 3.67E-28 |

| | | | | |
|---|---|---|---|---|
| The peak reference value for the lumbar spine (L3) | 90.13 (15.34) | 91.57 (16.19) | 89.88 (15.18) | 1.78E-03 |
| Age-matched value for lumbar spine (L3) | 98.44 (15.73) | 103.85 (17.07) | 97.53 (15.31) | 1.13E-27 |
| bone area in the lumbar spine (L4) | 16.80 (2.13) | 17.09 (2.23) | 16.75 (2.11) | 7.61E-06 |
| bone mineral content in the lumbar spine (L4) | 17.08 (4.08) | 17.81 (4.59) | 16.96 (3.97) | 3.62E-08 |
| bone mineral density in the lumbar spine (L4) | 1.01 (0.17) | 1.03 (0.18) | 1.01 (0.17) | 1.72E-05 |
| lumbar spine (L4) T-Score | -0.92 (1.52) | -0.73 (1.59) | -0.95 (1.50) | 4.22E-05 |
| lumbar spine (L4) Z-Score | -0.07 (1.47) | 0.46 (1.52) | -0.16 (1.44) | 4.30E-32 |
| The peak reference value for the lumbar spine (L4) | 90.85 (14.99) | 92.71 (15.67) | 90.54 (14.85) | 4.65E-05 |
| Age-matched value for lumbar spine (L4) | 99.38 (15.99) | 105.40 (17.17) | 98.38 (15.57) | 3.81E-32 |
| Spine total area | 57.58 (8.66) | 58.53 (9.91) | 57.42 (8.42) | 5.35E-04 |
| Spine total bone mineral content | 55.20 (14.41) | 57.75 (16.94) | 54.77 (13.89) | 5.90E-08 |
| Spine total bone mineral density | 0.95 (0.16) | 0.97 (0.17) | 0.95 (0.16) | 1.33E-06 |
| Spine total T-Score | -1.00 (1.42) | -0.85 (1.47) | -1.03 (1.41) | 1.70E-04 |
| Spine total Z-Score | -0.19 (1.33) | 0.29 (1.37) | -0.27 (1.30) | 2.93E-33 |
| Spine total peak reference value | 89.57 (14.78) | 91.16 (15.22) | 89.30 (14.69) | 2.18E-04 |
| Spine total age-matched value | 97.97 (15.15) | 103.44 (16.10) | 97.04 (14.79) | 2.65E-32 |
| Left hip k | 1.14 (0.00) | 1.14 (0.00) | 1.14 (0.00) | 1.81E-07 |
| Left hip d0 | 49.27 (1.48) | 48.93 (1.62) | 49.32 (1.44) | 2.49E-13 |
| The thickness of the left hip | 5.56 (0.55) | 5.69 (0.65) | 5.54 (0.53) | 1.90E-13 |
| Width of the region of interest in the left hip | 102.99 (7.14) | 104.65 (7.06) | 102.70 (7.12) | 2.45E-16 |
| Length of the region of interest in the left hip | 105.61 (8.62) | 106.43 (8.93) | 105.47 (8.56) | 1.17E-03 |
| The neck area of the left hip | 4.78 (0.56) | 4.86 (0.57) | 4.76 (0.56) | 1.24E-07 |
| Neckbone mineral of the left hip | 3.44 (0.76) | 3.54 (0.81) | 3.43 (0.75) | 4.02E-05 |
| Neckbone mineral density of the left hip | 0.72 (0.12) | 0.72 (0.12) | 0.72 (0.12) | 1.18E-01 |
| Neck T-Score of the left hip | -1.33 (0.98) | -1.36 (0.96) | -1.33 (0.98) | 3.88E-01 |
| Neck Z-Score of the left hip | -0.44 (0.90) | -0.16 (0.90) | -0.49 (0.90) | 2.98E-27 |
| Neck peak reference value of the left hip | 81.88 (13.20) | 81.35 (13.02) | 81.97 (13.23) | 1.51E-01 |
| Neck age-matched value of the left hip | 93.24 (14.01) | 97.40 (14.96) | 92.53 (13.72) | 1.93E-22 |
| Neck total area of the left hip | 34.88 (5.36) | 36.17 (5.51) | 34.66 (5.30) | 2.69E-16 |
| Neck total bone mineral of the left hip | 30.31 (7.87) | 32.49 (8.65) | 29.94 (7.67) | 6.66E-19 |
| Neck total bone mineral density of the left hip | 0.86 (0.13) | 0.89 (0.14) | 0.86 (0.13) | 1.35E-11 |
| Neck total T-Score of the left hip | -0.86 (0.96) | -0.75 (0.96) | -0.88 (0.96) | 1.28E-05 |
| Neck total Z-Score of the left hip | -0.29 (0.91) | 0.06 (0.91) | -0.34 (0.89) | 6.42E-38 |
| Neck total peak reference value of the left hip | 88.40 (12.80) | 89.89 (12.99) | 88.15 (12.75) | 5.58E-05 |
| Neck total age-matched value of the left hip | 95.85 (13.32) | 100.91 (14.12) | 94.99 (12.98) | 2.39E-35 |
| Right hip k | 1.14 (0.00) | 1.14 (0.00) | 1.14 (0.00) | 2.20E-12 |
| Right hip d0 | 49.33 (1.45) | 49.05 (1.56) | 49.38 (1.43) | 2.79E-10 |
| The thickness of the right hip | 5.53 (0.54) | 5.65 (0.63) | 5.51 (0.52) | 2.77E-12 |

| | | | | |
|---|---|---|---|---|
| Width of the region of interest in the right hip | 102.41 (7.01) | 104.33 (6.93) | 102.09 (6.97) | 1.01E-21 |
| Length of the region of interest in the right hip | 105.30 (8.61) | 106.10 (8.64) | 105.16 (8.60) | 1.13E-03 |
| Neck width of the right hip | 48.44 (1.47) | 48.51 (1.46) | 48.43 (1.47) | 8.27E-02 |
| The neck area of the right hip | 4.70 (0.57) | 4.81 (0.55) | 4.68 (0.58) | 1.90E-11 |
| Neckbone mineral of the Left Hip | 3.40 (0.76) | 3.54 (0.80) | 3.38 (0.75) | 7.61E-09 |
| Neckbone mineral density of the right hip | 0.72 (0.12) | 0.73 (0.12) | 0.72 (0.12) | 4.46E-03 |
| Neck T-Score of the right hip | -1.30 (0.98) | -1.29 (0.96) | -1.31 (0.98) | 6.24E-01 |
| Neck Z-Score of the right hip | -0.41 (0.91) | -0.09 (0.90) | -0.46 (0.90) | 1.40E-33 |
| Neck peak reference value of the right hip | 82.30 (13.23) | 82.26 (13.02) | 82.31 (13.27) | 9.00E-01 |
| Neck age-matched value of the right hip | 93.73 (14.07) | 98.47 (14.94) | 92.93 (13.76) | 2.61E-28 |
| Neck total area of the right hip | 34.57 (5.24) | 35.95 (5.25) | 34.34 (5.20) | 7.60E-20 |
| Neck total bone mineral of the right hip | 29.99 (7.68) | 32.32 (8.23) | 29.60 (7.51) | 3.87E-23 |
| Neck total bone mineral density of the right hip | 0.86 (0.13) | 0.89 (0.14) | 0.86 (0.13) | 1.11E-14 |
| Neck total T-Score of the right hip | -0.87 (0.94) | -0.73 (0.93) | -0.90 (0.95) | 7.22E-08 |
| Neck total Z-Score of the right hip | -0.30 (0.90) | 0.07 (0.89) | -0.36 (0.88) | 8.45E-45 |
| Neck total peak reference value of the right hip | 88.27 (12.64) | 90.07 (12.57) | 87.96 (12.63) | 5.26E-07 |
| Neck total age-matched value of the right hip | 95.70 (13.15) | 101.08 (13.73) | 94.80 (12.83) | 3.12E-41 |
| Dominant limb for bone density measurement | 148 (2.0%) | 26 (2.4%) | 122 (1.9%) | 4.72E-01 |
| Bone Stiffness Index | 90.17 (17.42) | 87.55 (16.31) | 90.61 (17.56) | 2.59E-08 |
| Percentile in young adults | 94.03 (18.00) | 89.39 (15.37) | 94.82 (18.29) | 2.72E-24 |
| T-SCORE | -0.53 (1.62) | -0.95 (1.37) | -0.46 (1.65) | 1.85E-24 |
| Percentile in age-matched adults | 111.92 (19.41) | 113.53 (19.03) | 111.64 (19.46) | 2.92E-03 |
| Z-SCORE | 0.90 (1.47) | 0.94 (1.33) | 0.89 (1.49) | 2.75E-01 |

**Table S2. Comparison between prediction models using default parameters and tuned parameters.**

A grid search method was applied to optimize several parameters in the best model ( $ABD_{GNN}^{PIX} + BMD_{COMBINE}^{PIX} + G + D$ ). The tuned parameters encompassed the learning rate (0.001, 0.01, 0.3), the minimum loss reduction required for splitting (gamma = 0, 0.1, 1, 1.5), the maximum depth of a tree (max_depth = 3, 6, 9), the minimum sum of instance weight required in a child (min_child_weight = 1, 5, 10), the subsample ratio of training instances (subsample = 0.8, 0.9, 1), and the control of the balance between positive and negative weights (scale_pos_weight = 0, 1, 2, 4). The default parameters were set as follows: learning rate = 0.3, gamma = 0, max_depth = 6, min_child_weight = 1, subsample = 1, and scale_pos_weight = 1. Out of the 1,296 training combinations, the optimal parameters were determined to be: learning rate = 0.3, gamma = 0, max_depth = 3, min_child_weight = 1, subsample = 0.8, and scale_pos_weight = 2.

| Data | Parameter | Performance | | | | |
|---|---|---|---|---|---|---|
| | | AUC | Accuracy | Sensitivity | Specificity | Youden |
| Testing data 1 | Default | 0.944 | 0.868 | 0.889 | 0.865 | 0.754 |
| | Tuned | 0.935 | 0.842 | 0.889 | 0.834 | 0.723 |
| Testing data 2 | Default | 0.954 | 0.875 | 0.882 | 0.875 | 0.757 |
| | Tuned | 0.942 | 0.851 | 0.902 | 0.844 | 0.746 |

### Supplementary Texts

**Supplemental Text 1: BMD image pre-processing.**

After cropping, the image size was not identical. For the spine image, the image height was 250 or 251, and the width range was 63 to 389. The image height for the left hip image was 241, 250, or 251, and the width range was 155 to 299. For the right hip image, the image height was 236, 241, 250, or 251, and the width range was 161 to 299. We used padding to make the image size consistent to 250 x 389, 250 x 299, and 250 x 299 in the spine, left hip, and right hip, respectively.
